# Anti-EPCR autoantibodies in ulcerative colitis show sex- and lipid-dependent patterns

**DOI:** 10.1101/2025.10.18.25337568

**Authors:** Nerea Ugidos-Damboriena, Llùcia Jaime-Gómez, Cristina Rodríguez-Gutiérrez, Lucía Zabalza, Óscar Nantes-Castillejo, María Gilda Dichiara-Rodríguez, Jacinto López-Sagaseta

## Abstract

Autoantibodies targeting the endothelial protein C receptor (EPCR) have been associated with ulcerative colitis (UC). We aimed to assess a potential role of EPCR lipidation on the detection of anti-EPCR antibodies in inflammatory bowel disease (IBD). To this end, serum samples from patients with UC, Crohn’s disease, and healthy controls were analyzed using an in-house ELISA employing either native or delipidated EPCR. Overall, male UC patients showed significantly higher absorbance (mean=1.09) than CD patients (mean=0.50) and controls (mean=0.37). Remarkably, in males, the difference between UC patients and controls was highly significant (p=1.2e-07), as was the difference between UC and CD patients (p=5.7e-05). In women, the difference was less pronounced and only significant when comparing UC to controls (p=0.023). Replacement of native EPCR with delipidated EPCR in the ELISA procedure dropped detection and eliminated the UC-CD discrimination power (p=0.784). These findings indicate a role for the bound lipid as key determinant of male-biased, anti-EPCR reactivity, and support the diagnostic potential of this biomarker when assay conditions preserve the physiological lipid-bound state of EPCR.

## Results and discussion

The presence of high anti-EPCR autoantibody titres has emerged as a potential biomarker in varied autoimmune diseases^1,2^. EPCR is pivotal for the integrity of the endothelium and plays critical roles in the anticoagulation system^3^. Moreover, abrogation of EPCR’s functions leads to a pro-inflammatory state, with a particular impact on chronic autoimmune disorders such as inflammatory bowel disease (IBD), which includes ulcerative colitis (UC) and Crohn’s disease (CD). Built on previous work by Mutoh *et al*^*4*^, which represents the first association of EPCR as a novel autoantigen in UC, a recent study by Kakuta and colleagues^5^ claims the diagnostic potential of anti-EPCR autoantibodies in UC. The authors demonstrated that anti-EPCR titres are significantly increased in UC patients compared to both healthy individuals and those with other colorectal disorders. They also found a high anti-EPCR prevalence in patients with left-sided or extensive UC, and those presenting UC comorbidities, such as arthritis and erythema nodosum. Further, Sawahashi *et al*^*6*^ performed a large prospective cohort, showing that anti-EPCR antibodies (alone or combined with anti-αvβ6) can predict UC years before clinical onset. Together, these results provide a solid ground for EPCR autoantibodies as promising biomarkers in IBD. Here, by using an in house-developed ELISA assay **(Figure 1A)**, we interrogated any sex bias associated with anti-EPCR levels, but also, a potential mechanistic basis underlying the pathogenic recognition of EPCR. Three cohorts were established by collecting blood samples from healthy subjects (controls) and patients with UC and CD **(Supplementary Table 1)**. Analysis of anti-EPCR levels across the different groups indicate that the median absorbance values in the UC patients is notably higher than in CD and controls **(Figure 1B)**. This preliminary observation aligns with previous reports^4–6^. The interquartile range is wider in the UC cohort, suggesting stronger variability in the anti-EPCR levels. Both disease type (p=0.028) and sex (p=0.018) exhibited a notable impact on anti-EPCR autoantibody levels. A strong interaction was also noticed between disease type and sex (p=0.024), suggesting that disease status influences anti-EPCR titres differently in males and females. Overall, male UC patients showed significantly higher absorbance (mean=1.09) than CD patients (mean=0.50) and controls (mean=0.37) **(Figure 1C and Supplementary Table 2)**. Remarkably, in males, the difference between UC patients and controls was highly significant (p=1.2×10^-7^), as was the difference between UC and CD patients (p=5.7×10^-5^). In women, while UC patients showed higher absorbance (mean=0.71) than controls (mean=0.45), the difference was less pronounced and only significant when comparing UC to controls (p=0.023). Comparison of CD and controls led to no significant differences. Thus, these observations indicate a strong and sex-specific immune response characterized by elevated anti-EPCR autoantibody levels in males with UC. Furthermore, the poor increase of anti-EPCR antibody levels in CD patients supports that the determination of anti-EPCR autoantibody titres is a promising biomarker to discriminate between UC and CD, particularly in males.

**Figure 1.**
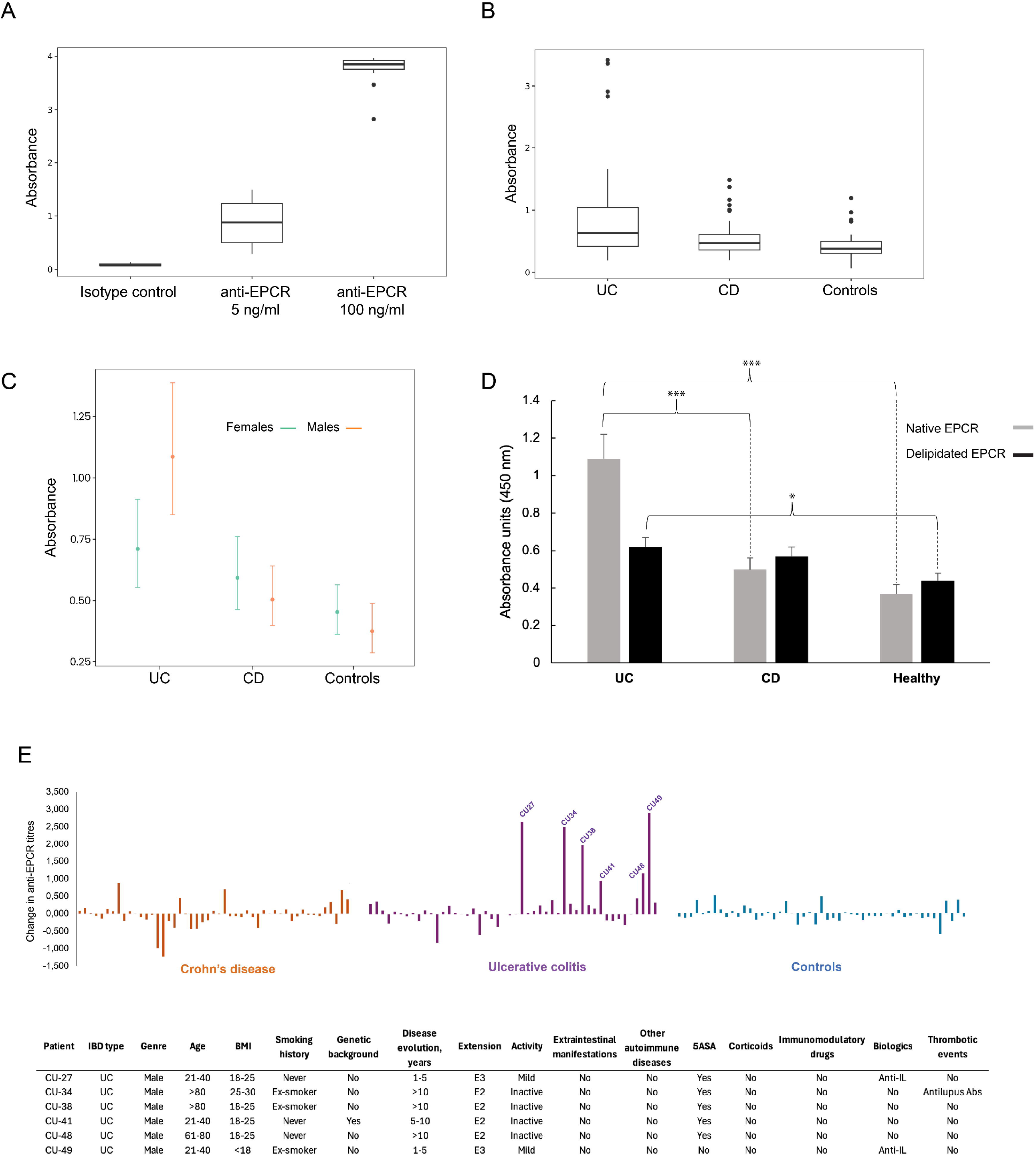
Differences in EPCR reactivity by disease type, sex and antigen lipidation. (A) The specificity of the ELISA was assessed with a dose-dependent binding response to commercial, recombinant anti-EPCR (5 ng/ml and 100 ng/ml) compared with isotype control. (B) Box plots displaying anti-EPCR absorbance levels (450 nm) in samples from patients suffering of ulcerative colitis (UC), Crohn’s disease (CD), and healthy controls. (C) Plots showing mean absorbance values ± error bars (95% CI) stratified by sex (females vs males) within each cohort. (D) Differences observed in anti-EPCR absorbance values in males as determined with lipidated (gray) and delipidated (black) EPCR in UC, CD, and controls. The y-axis represents absorbance units (450 nm). Statistical significance between groups: *p < 0*.*001: ***; p < 0*.*01: **; p < 0*.*05: *; p < 0*.*1*. (E) Differences in anti-EPCR titres were calculated by subtracting the values obtained with delipidated EPCR from those obtained with native EPCR. The bar plots highlight the patients with the highest difference values, all of whom fall within the UC cohort. The table below details clinical characteristics of these patients.

EPCR is a transmembrane protein characterized by the presence of a bound phospholipid in a CD1-like groove that shapes the receptor’s structure^7,8^. The lipid hydrophobic region is buried in this pocket, while the polar head group is exposed to the outside, being potentially accessible for molecular recognition^9,10^. Thus, we explored whether the anti-EPCR antibodies could target, to a greater or lesser extent, the bound lipid. To this end, we analysed the patient samples using delipidated EPCR. Under this modified protocol, and relative to controls, both UC and CD patients still exhibited increased levels of anti-EPCR autoantibodies. However, while the UC vs. control comparison remained statistically significant (p=0.014) **(Figure 1D and Supplementary Table 2)**, supporting the robustness of the UC-associated signal, the modified ELISA yield lowered anti-EPCR titres, particularly in UC males (1.09 vs 0.62), resulting in a reduced stratification between UC and CD patients. CD patients showed a borderline elevation compared to controls (p=0.077), indicating that the altered assay may be more sensitive to lower-level responses or potentially less specific. However, the discrepancy between the anti-EPCR levels in UC and CD patients was no longer significant (p=0.784), suggesting that the new assay had a reduced discriminatory power.

By comparing the differences in anti-EPCR levels when using the ELISA assay with native or delipidated EPCR, we detected several high outliers that highlight a subset of UC patients with strong reactivity towards the lipid moiety **(Figure 1E)**. These patients are characterized by being all males with UC, either left-sided colitis (E2) or extensive colitis (E3), consistent with broader epithelial disruption and possibly more systemic immune exposure to EPCR. The fact that these patients show mild to negligible activity suggest they are not an active signal of inflammation, or that they reflect immune imprinting from previous mucosal injury or inflammation.

Overall, these observations suggest that a significant fraction of the anti-EPCR autoantibodies are directed against the bound lipid or a motif that involves both a fraction of EPCR’s and its bound lipid. Removal or alteration of the bound lipid results in a potential loss in diagnostic accuracy, particularly in distinguishing UC from CD, and with a significant relevance in males. The drop in the discriminatory power underscores the importance of the assay design in preserving clinically and mechanistically relevant motifs that are critical for recognition by circulating anti-EPCR antibodies.

Our findings add two elements that had not yet been explored in the context of anti-EPCR levels in IBD. On the one hand, this study provides evidence for a role of the bound lipid in driving antigen recognition by a significant fraction of the anti-EPCR antibodies found in UC patients. On the other hand, we observed a statistically higher proportion of anti-EPCR antibodies in males with UC compared to females. These findings unlock features relevant for the clinical application of these antibodies in IBD diagnostics. Moreover, this study encourages further research to address the question of whether this lipid-driven reactivity associates with a specific gut microbiota that underlies UC.

## Acknowledgements

We thank Sergio Morales-Hernández for technical contributions. We also thank Eduardo Albéniz for initial discussions concerning the focus of the study.

## Author contributions

Conceptualization: JLS; Methodology: NUD, LLJG, CRG, LZ, ON, MGDR and JLS; Manuscript writing: JLS.

## Institutional Review Board Statement

This study was carried out according to the Declaration of Helsinki and was approved by the Drug Research Ethics Committee of Navarra (PI_2021/11).

## Informed Consent Statement

Informed consent was obtained from all subjects involved in the study.

## Data availability

Due to an active patent application, proprietary details regarding of the ELISA protocol are not disclosed in this preprint. Additional technical information is available from the corresponding author (J.L.S.) upon reasonable request for academic, clinical, and non-commercial use.

## Supplementary Material

**Supplementary Table 1.** Characteristics of the three cohorts analyzed in this study.

| Characteristic | UC (n = 48) | CD (n = 48 ) | Controls (n = 50) |
| --- | --- | --- | --- |
| **Age range** |  |  |  |
| <20 | 0 | 2 | 1 |
| 21-40 | 17 | 14 | 17 |
| 41-60 | 20 | 24 | 24 |
| 61-80 | 8 | 8 | 8 |
| >80 | 3 | 0 | 0 |
| **Females/males (%):** | 51/49 | 46/54 | 58/42 |
| **IMC range (%)** |  |  |  |
| <18 | 4.2 | 4.3 | 0 |
| 18-25 | 62.5 | 55.3 | 63.4 |
| 25-30 | 25.0 | 29.8 | 29.3 |
| 30-40 | 6.2 | 10.6 | 7.3 |
| >40 | 2.1 | 0 | 0 |
| **Smoking habits (%)** |  |  |  |
| Never | 56 | 36 | 74 |
| Ex | 34 | 28 | 16 |
| smoker | 10 | 36 | 10 |
| **Extension (%)** |  |  |  |
|  | (E1) 10.4 | (L1) 42.0 | does not apply |
|  | (E2) 39.6 | (L1 L4) 12.0 | does not apply |
|  | (E2/E3) 2.1 | (L2) 4.0 | does not apply |
|  | (E3) 43.8 | (L2 L4) 2.0 | does not apply |
|  | (E4) 2.1 | (L3) 24.0 | does not apply |
|  | (E5) 2.1 | (L3 L4) 14.0 | does not apply |
|  |  | (L4) 2.0 | does not apply |
| **Disease lifetime, years (%)** |  |  |  |
| <1 | 6.0 | 6.0 | does not apply |
| 1-5 | 20.0 | 22.0 | does not apply |
| 5-10 | 18.0 | 12.0 | does not apply |
| >10 | 56.0 | 60.0 | does not apply |
| **Disease activity (%)** |  |  |  |
| Inactive | 81.6 | 54.0 | does not apply |
| Mild | 14.3 | 18.0 | does not apply |
| Moderate | 4.1 | 28.0 | does not apply |
| **Lipid-dependent anti-EPCR levels (%)** | 70.8 | 42.9 | 32.7 |

**Supplementary Table 2.** Effect of EPCR lipidation on ELISA absorbance. Preservation of EPCR lipidation (Native EPCR) leads to higher absorbance values and UC/CD stratification only in males. In contrast, delipidated EPCR results in a reduced detection power and loss of disease discrimination. The upper values (Native EPCR) indicate male-only data, while the values in the lower section (Delipidated EPCR) applies to all patients. Significant results are indicated with asterisks: p < 0.001: ***; p < 0.01: **; p < 0.05: *; p < 0.1; UC: Ulcerative colitis; CD: Crohn’s disease.

| Native EPCR |  |  |  |
| --- | --- | --- | --- |
| DISEASE | Emmean (CI 95%) | contrast | p.value |
| UC | 1.09 ± 0.13 [0.85 - 1.39] | UC/CD | 5.7e-05*** |
| CD | 0.50 ± 0.06 [0.4 - 0.64] | UC/Control | 1.2e-07*** |
| Controls | 0.37 ± 0.05 [0.29 - 0.49] | CD/Control | 0.230 |
| Delipidated EPCR |  |  |  |
| UC | 0.62 ± 0.05 [0.53 - 0.73] | Control/UC | 0.014* |
| CD | 0.57 ± 0.05 [0.49 - 0.67] | Control/CD | 0.077 |
| Controls | 0.44 ± 0.04 [0.38 - 0.52] | UC/CD | 0.784 |

